# Cortical surface area, thalamic nuclei volume, and associations with neurocognitive functioning in 6–13-year-old children born late preterm and full term

**DOI:** 10.64898/2026.09.15.26362093

**Authors:** Paige M. Nelson, Heidi M. Harmon, Jane E. Brumbaugh, Peggy C. Nopoulos, Jerome J. Maller, Allison M. Momany, Vincent A. Magnotta

## Abstract

**Background:** Late preterm (LP) children (34 0/7 – 36 6/7 weeks of gestation) are at risk for neurocognitive challenges, yet the neuroanatomic and functional disruptions underlying these developmental differences remain poorly understood.

**Purpose or Hypothesis:** Investigate (1) regional differences in cortical surface area and thalamic nuclei volume and (2) structural variations that may explain heterogeneity in neurocognitive functioning in LP versus full term (FT) school-aged children, using two complementary approaches: a vertex-wise analysis of cortical surface area and a region-of-interest volumetric analysis of thalamic nuclei.

**Study Type:** Secondary analysis of a prospective observational cohort study.

**Population or Subjects:** 48 LP and 72 FT children aged 6–13 years.

**Field Strength/Sequence:** 3T; Sagittal T1-weighted MP-RAGE and axial T2-weighted 3D SPACE.

**Assessment:** FreeSurfer 7.4.1 was used to parcellate cortical regions (Destrieux atlas) and segment thalamic nuclei (25 nuclei). Neurocognitive functioning was assessed across four domains: hyperactivity/inattention (Pediatric Behavior Scale), processing speed (WISC-IV), working memory (WISC-III), and visual-spatial perception (Benton JLO).

**Statistical Tests:** Independent samples t-tests for group comparisons; vertex-wise GLMs for cortical surface analyses with false discovery rate (FDR) correction; and multiple linear regression for thalamic nuclei analyses with FDR-corrected *q* < 0.05.

**Results:** Vertex-wise analyses revealed regional differences in cortical surface area between LP and FT children in both hemispheres, along with associations between surface area in several cortical regions and hyperactivity/inattention, working memory, and visual-spatial perception. In contrast, using an ROI approach, thalamic nuclei volumes did not differ between groups and were not associated with any neurocognitive domains.

**Conclusion:** LP birth may be associated with subtle, region-specific bidirectional left and right hemispheric cortical surface area differences rather than widespread structural alterations in middle childhood, with regional macrostructural measures associated with individual differences in neurocognitive functioning.

## Introduction

Each year, approximately 15 million newborns,^1^ or about 12% of newborns worldwide, are born preterm (< 37 0/7 weeks of gestation).^2^ Historically, much of the research on development outcomes to date has been focused on neonates born less than 34 0/7 weeks of gestation,^3,4^ despite late preterm (LP) birth (34 0/7–36 6/7 weeks of gestation) representing nearly 80% of all preterm deliveries in the United States.^5^ Notably, LP neonates demonstrate metabolic, neurologic, and physiologic immaturities similar to that observed in preterm children born earlier, only less severe.^6^ Compared with neonates born at full term (FT; ≥ 37 0/7 weeks of gestation), LP neonates often face higher risk of neonatal morbidities (e.g., respiratory distress syndrome, hypoglycemia, feeding difficulties),^7^ mortality,^8,9^ neonatal hospital readmission,^10,11^ and create greater familial and societal financial burden^12,13^ – all of which are factors that may collectively influence developmental trajectories.

Recent investigations suggest that LP children are at risk of short- and long-term neurocognitive, socioemotional/behavioral, and psychiatric challenges compared to their FT counterparts.^14–16^ A systematic review found higher rates of neurodevelopmental disabilities, lower educational achievement, increased need for early intervention services, more medical disabilities (e.g., visual impairment/blindness; hearing impairment/deafness; and seizure disorders), and poor physical growth among LP children aged 1 to 7 years compared to their FT peers.^17^ Additionally, a recent nationwide cohort study demonstrated that LP children had an increased risk of cognitive, motor, epileptic, visual, and hearing impairments compared with their FT counterparts, with outcomes followed through age 16 years (median follow-up, 13.1 years; interquartile range, 9.5–15.9 years).^5^ Yet, it remains unclear why some LP children achieve favorable short-term and long-term developmental sequelae while others experience significant challenges,^18–20^ as the neuroanatomic and functional disruptions underlying these developmental differences remain poorly understood.

The immature brain is highly vulnerable to perinatal brain insults and dysmaturation during the gestational window of 34 to 40 weeks; this phase is one of the most dynamic and influential phases of in-utero neurodevelopment, where cortical volume increases by 50% and 25% of cerebellar development occurs.^21–24^ The premature transition of the immature brain to the extrauterine environment disrupts normal third trimester neurodevelopmental processes (e.g., axonal elongation, synaptogenesis, and myelination),^25^ and can lead to diffuse white matter injury, altered cortical development, reduced myelination, and disrupted structural connectivity.^26–29^ Markedly, studies of LP neonates have revealed neuropathological alterations even in the absence of macroscopic injuries, as demonstrated by reduced size of the corpus callosum, deep grey nuclei, and cerebellum,^30^ as well as delayed myelination of the posterior limb of the internal capsule, immature gyral maturation, and increased cerebrospinal fluid spaces at approximately 36 weeks postmenstrual age compared to FT neonates.^31^ Likewise, reduced cortical surface area and complexity have been exhibited around term-equivalent age, or 38–42 weeks postmenstrual age, in preterm neonates born less than 30 weeks of gestation, as compared to FT neonates, despite similar cerebral tissue volumes.^32^

Longitudinal volumetric MRI evidence has demonstrated that structural brain differences in very preterm children (< 32 0/7 weeks of gestation) persist and evolve throughout late childhood and early adolescence, with one study documenting attenuated grey matter growth and reduced white matter gain between 8 and 12 years of age in children born weighing less than 1250 grams, as compared to FT counterparts.^33^ Similarly, Rathbone et al. 2011 found that decreases in cortical surface area growth, rather than total brain volume growth, were associated with a 1 standard deviation fall in global neurocognitive scores (5 –11% lower surface area per standard deviation) assessed at two and six years, particularly in attention, planning, memory, language, and numeric and conceptual abilities, with no such link detected for motor skills.^34^ Thus, it remains critical to examine whether the neuroanatomical differences observed in LP neonates persist into middle childhood – an important and developmentally meaningful window – and whether they can help elucidate the discrepant neurocognitive outcomes in LP children.

Our team previously conducted an observational study evaluating differences in structural brain measures, as well as cognitive and behavioral outcomes, between LP and FT school-aged children (6–13 year old).^35^ Findings showed that LP children exhibited reduced total brain tissue volume, cerebral volume, and cortical surface area; greater cerebrospinal fluid volume; and a relatively smaller thalamus, while other subcortical structures (hippocampus, caudate, putamen, and globus pallidus) did not differ significantly. In addition, LP children demonstrated significant deficits in processing speed, visual-spatial perception, and working memory, in addition to greater parent-reported behavioral difficulties (hyperactivity/inattention), as compared to their FT counterparts. While these findings established that LP birth has lasting effects on both macroscopic brain measures and neurocognitive function, the global nature of the structural measures – total surface area and whole thalamic volume – masked determination of whether the underlying differences were regionally specific or diffusely distributed across the cortical surface and thalamic nuclei.

Thus, in this study, we aimed to move beyond characterizing LP brain differences as global reduction in cortical surface area and thalamic volume and instead identify which specific cortical regions and thalamic nuclei are affected and, critically, whether those regional differences explain the neurocognitive deficits observed in this population. Specifically, we combined a vertex-wise analysis of cortical surface area with a region-of-interest (ROI) volumetric analysis of thalamic nuclei to investigate (1) whether there are regional differences in cortical surface area and thalamic nuclei volumetrics between LP and FT children and (2) whether structural variations, more broadly, help explain heterogeneity in neurocognitive functioning (hyperactivity/inattention, processing speed, working memory, and visual-spatial perception) in children aged 6 to 13 years, while accounting for gestational age group (LP and FT).

## Methods

### Study Population

Secondary analysis was conducted on a sample of 52 LP (34 0/7 to 36 6/7 weeks of gestation) and 74 FT (≥ 37 0/7 weeks of gestation) children, ages 6 to 13 years, who were born between 1996 and 2006. Exclusion criteria were as follows: 5-minute Apgar score <7, birth weight <1500 grams for LP children or <2500 grams for FT children, multiple births, major chromosomal anomalies or congenital malformations, major neonatal morbidities (e.g., sepsis), neonatal encephalopathy, neonatal seizures (excluding febrile seizures), brain tumors or prior brain surgery, and/or uncorrected hearing loss. During the study visit, children completed comprehensive neurocognitive assessments and had a structural MRI. Children with incomplete neuroimaging data were excluded (motion artifacts for 3 LP children; scan sequences stopped early or not attempted for one LP, two FT children), resulting in usable structural MRI data for 48 LP and 72 FT children. This study was approved by the Institutional Review Board and written parental consent and child assent (written for ages 8–13, verbal for ages 6–7) were obtained. Comprehensive details regarding study design, recruitment procedures, neuroimaging protocols, and neurocognitive assessments are available in Brumbaugh et al. 2016.^35^

### Neurocognitive Assessments

Neurocognitive functioning was assessed across four domains – attention, processing speed, visual-spatial perception, and working memory – based on previous findings in this cohort showing significant differences between LP and FT children (see Brumbaugh et al. 2016 for full battery and procedures).^35^ The parent-reported Pediatric Behavior Scale (PBS) was used to derive a scaled score of *hyperactivity/inattention*.^36^ Processing speed, working memory, and visual-spatial perception were assessed by clinician-administered standardized neurocognitive assessments. Specifically, *processing speed* was assessed using the Processing Speed Index (composite score) from the Wechsler Intelligence Scale for Children, Fourth Edition (WISC-IV),^37^ *working memory* was assessed using the Spatial Span subtest (scaled score) from the Wechsler Intelligence Scale for Children, Third Edition (WISC-III),^38^ and *visual-spatial perception* was evaluated via the Benton Judgment of Line Orientation (JLO; z-score).^39^

### Brain MRI Acquisition

Structural neuroimaging was performed without sedation in a Siemens 3T TIM Trio scanner (Siemens Healthcare, Erlangen, Germany) using a 12-channel head coil (Brumbaugh et al. 201635 for detailed scanning protocol). T1-weighted images were acquired using a sagittal MP-RAGE sequence (TR=2300ms; TE=2.82ms; TI=1100ms; flip angle=10°; FOV=180 × 180 × 224 mm; matrix=256 × 256 × 240; bandwidth 200 Hz/pixel), followed by axial T2-weighted images collected with a 3D SPACE sequence (TR=9910ms; TE=430ms; FOV=180 × 180 × 192 mm; matrix=256 × 256 × 128).

### Cortical Surface Area and Thalamic Nuclei Volumetric Processing

Cortical reconstruction and volumetric segmentation using the T1- and T2-weighted scans were performed using FreeSurfer v.7.4.1 (http://surfer.nmr.mgh.harvard.edu/). The resulting cortical surface models were visually inspected, and automated quality control was completed using Qoala-T v1.2, a supervised learning tool designed to assess pediatric FreeSurfer-processed scans (http://github.com/Qoala-T/QC; Klapwijk et al. 2019).^40^

For vertex-wise analysis of cortical surface area, individual surface reconstructions were registered to the *fsaverage* template, parcellated, and anatomically labeled using the Destrieux cortical atlas (aparc.a2009s) which identifies 148 cortical regions (74 per hemisphere),^41^ and smoothed using a 10mm FWHM Gaussian kernel, implemented via the FreeSurfer module QDEC.

In addition, thalamic nuclei volumes were derived according to the probabilistic atlas described by Iglesias et al. 2018.^42^ Thalamic nuclei volumes were obtained separately for the left and right hemispheres and subsequently normalized using the estimated total intracranial volume (eTIV) to account for individual differences in head size.

### Statistical Analysis

Descriptive statistics were computed for demographics, birth characteristics, and anthropometrics. Bivariate comparisons between LP and FT children were conducted using chi-square tests of independence (χ^2^) for categorical variables and independent samples t-tests for continuous variables.

For cortical surface area, vertex-wise general linear models (GLMs) were estimated using QDEC; first, a GLM was estimated at each vertex to test for group differences in cortical surface area between FT and LP children (i.e. contrast was FT-LP), covarying for total brain volume (TBV). Second, separate vertex-wise GLMs were estimated to examine associations between cortical surface area and each neurocognitive outcome, accounting for gestational age group (FT versus LP). Correction for multiple comparison across the cortical surface was performed using the false discovery rate (FDR) method, with significance set at FDR-corrected *q* < 0.05.

To assess associations between thalamic nuclei and neurocognitive outcomes, a series of multiple linear regressions were conducted in R v4.4.3 utilizing the *lm* function from the *stats* package (R Core Team, 2025). Each neurocognitive outcome was modeled as a dependent variable, with normalized thalamic nuclei volumes as independent variables. Given the number of thalamic nuclei and neurocognitive outcomes analyzed, *p*-values were adjusted for multiple comparisons using the FDR method to control for Type I error; significance was set at an FDR-adjusted *q* < 0.05.

## Results

### Participant Characteristics

Age, maternal educational achievement, biological sex at birth, weight, and occipitofrontal circumference at the time of the brain MRI and neurocognitive testing were similar between LP and FT children (all *p-*values > 0.05). LP children were more likely to have lower birth weights (2.6 kg versus 3.6 kg, *p* < 0.001), and at the time they underwent brain MRI and neurocognitive testing, they were also significantly shorter (136.5 cm versus 143.5 cm, *p* = 0.009; Table 1).

**Table 1.** Demographics and anthropometrics between FT and LP children.

| | Gestational Age | | $p$-value |
| --- | --- | --- | --- |
|  | Full Term<br>(n=72) | Late Preterm<br>(n=48) |  |
| Birth weight, *kg* | 3.6 ± 0.4 | 2.6 ± 0.6 | < 0.001 |
| Age at MRI/neurocognitive testing, *years* | 10.0 ± 2.2 | 9.6 ± 1.9 | 0.243 |
| Maternal educational achievement, *years* | 16.4 ± 2.0 | 15.7 ± 2.3 | 0.086 |
| Male, *n (%)* | 35 (49%) | 26 (54%) | 0.681 |
| Height, *cm* | 143.5 ± 14.3 | 136.5 ± 13.8 | 0.009 |
| Weight, *kg* | 38.9 ± 12.7 | 35.2 ± 14.5 | 0.143 |
| Occipitofrontal circumference, *cm* | 55.2 ± 4.4 | 53.9 ± 2.9 | 0.083 |

### Vertex-Wise Analysis of Cortical Surface Area Across Gestational Age and Neurocognitive Functioning

LP children exhibited significantly greater (blue regions) cortical surface area in the left hemisphere in one region (lateral orbitofrontal; Figure 1A) and right hemisphere in four regions (lateral occipital, pars opercularis, inferior temporal, and precentral; Figure 1B) when controlling for TBV in the model (FDR *q* < 0.05; Table 2). In addition, LP children exhibited significant reduction (orange region) in cortical surface area relative to FT children in the right hemisphere in one cluster (banks of superior temporal sulcus; Figure 1B) in the same statistical model (FDR *q* < 0.05; Table 2). No other significant differences emerged between the groups in the vertex-wise analysis of the cortical surface area after FDR correction for multiple comparisons.

**Table 2.** Left and right hemisphere cortical surface area clusters showing significant vertex-wise differences between full term and late preterm children, controlling for total brain volume.

| Region | Max Statistic | Cluster Size (mm<sup>2</sup>) | Tal X | Tal Y | Tal Z | *p*-value | 95% CI Low | 95% CI High | # Vertices |
| --- | --- | --- | --- | --- | --- | --- | --- | --- | --- |
| LH Lateral Orbitofrontal | -3.1862 | 1433.52 | -27.1 | 26.3 | 2.4 | 0.03220 | 0.0299 | 0.0345 | 2912 |
| RH Banks of Superior Temporal Sulcus | 3.2692 | 2221.33 | 45.0 | -39.8 | 12.3 | 0.00180 | 0.0013 | 0.0024 | 5358 |
| RH Lateral Occipital | -3.3338 | 2192.88 | 27.8 | -85.7 | 14.6 | 0.00190 | 0.0014 | 0.0025 | 3092 |
| RH Pars Opercularis | -4.4086 | 1967.02 | 38.8 | 11.8 | 9.6 | 0.00440 | 0.0036 | 0.0053 | 4339 |
| RH Inferior Temporal | -2.8309 | 1423.39 | 51.8 | -17.9 | -29.0 | 0.03450 | 0.0322 | 0.0368 | 2259 |
| RH Precentral | -2.3848 | 1416.33 | 35.3 | -4.6 | 43.9 | 0.03580 | 0.0334 | 0.0382 | 2900 |

*Abbreviations.* LH, left hemisphere; RH, right hemisphere.

**Figure 1.**
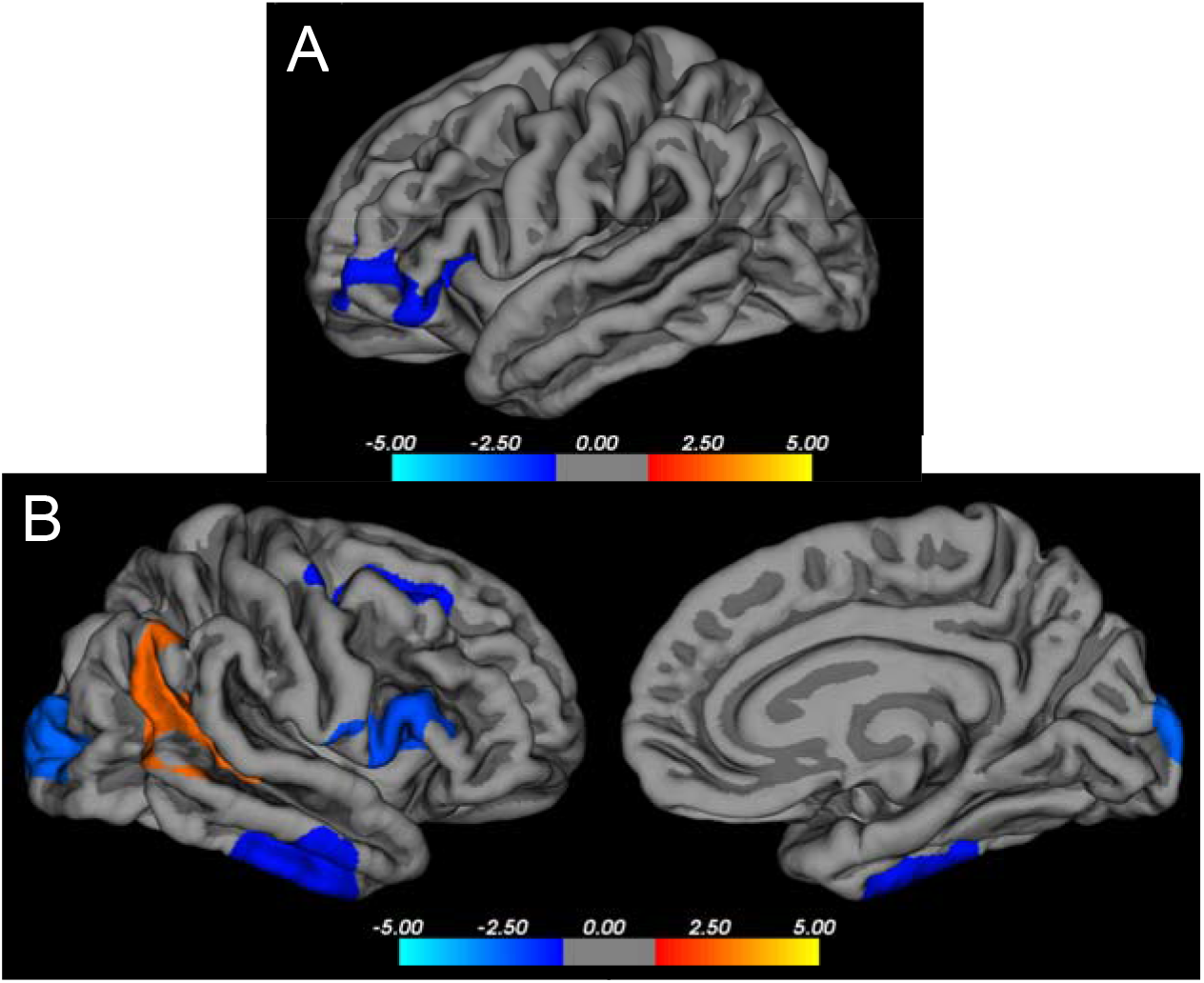
Vertex-wise comparison of left (A) and right (B) hemispheric cortical surface area between groups. Difference between full term (FT) and late preterm (LP) children (FT-LP) controlling for total brain volume, corrected for FDR *q* < .05, on the FreeSurfer average surface. Blue represents regions where LP surface areas are larger than FT, and orange indicates regions where LP surface areas are significantly smaller than FT.

Furthermore, when assessing cortical surface area in relation to neurocognitive outcomes, the cortical surface area of the right caudal middle frontal region was negatively correlated with hyperactivity/inattention, whereby less surface area was associated with increased scores on the PBS Hyperactivity/Inattention subscale, after accounting for gestational age group (LP versus FT) (FDR-corrected *q* < 0.05; Figure 2; Table 3).

**Table 3.**
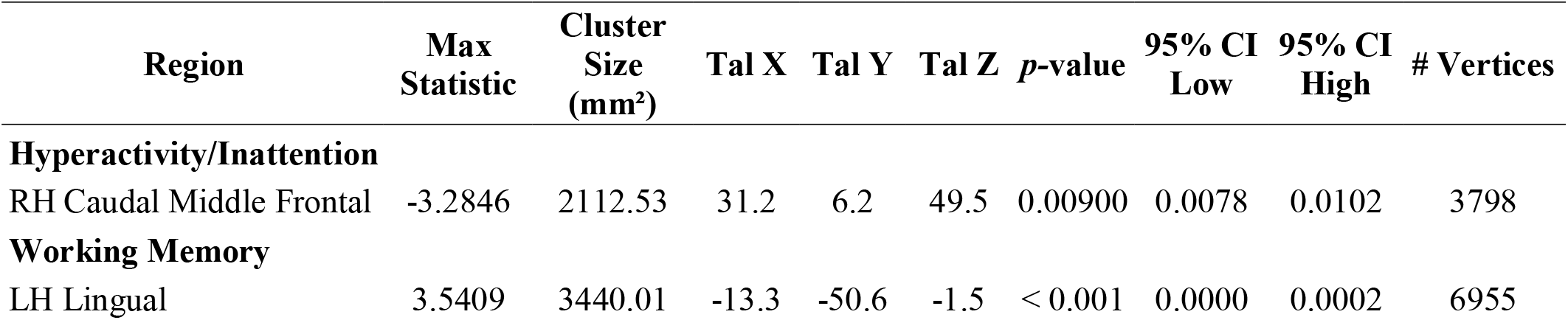
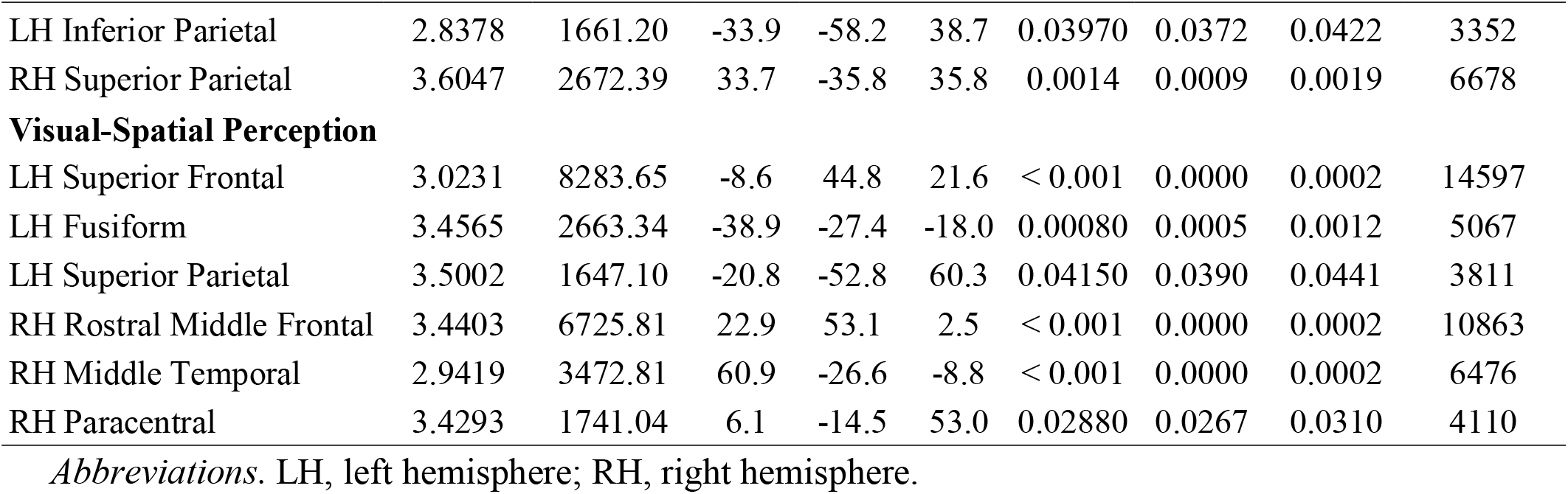
Left and right hemisphere cortical surface area clusters showing significant associations with neurocognitive domains, accounting for gestational age group.

| Region | Max Statistic | Cluster Size (mm<sup>2</sup>) | Tal X | Tal Y | Tal Z | p-value | 95% CI Low | 95% CI High | # Vertices |
| --- | --- | --- | --- | --- | --- | --- | --- | --- | --- |
| **Hyperactivity/Inattention** |  |  |  |  |  |  |  |  |  |
| RH Caudal Middle Frontal | -3.2846 | 2112.53 | 31.2 | 6.2 | 49.5 | 0.00900 | 0.0078 | 0.0102 | 3798 |
| **Working Memory** |  |  |  |  |  |  |  |  |  |
| LH Lingual | 3.5409 | 3440.01 | -13.3 | -50.6 | -1.5 | < 0.001 | 0.0000 | 0.0002 | 6955 |

| Region | Max<br>Statistic | Cluster<br>Size<br>(mm<sup>2</sup>) | Tal X | Tal Y | Tal Z | p-value | 95% CI<br>Low | 95% CI<br>High | # Vertices |
| --- | --- | --- | --- | --- | --- | --- | --- | --- | --- |
| LH Inferior Parietal | 2.8378 | 1661.20 | -33.9 | -58.2 | 38.7 | 0.03970 | 0.0372 | 0.0422 | 3352 |
| RH Superior Parietal | 3.6047 | 2672.39 | 33.7 | -35.8 | 35.8 | 0.0014 | 0.0009 | 0.0019 | 6678 |
| **Visual-Spatial Perception** |  |  |  |  |  |  |  |  |  |
| LH Superior Frontal | 3.0231 | 8283.65 | -8.6 | 44.8 | 21.6 | < 0.001 | 0.0000 | 0.0002 | 14597 |
| LH Fusiform | 3.4565 | 2663.34 | -38.9 | -27.4 | -18.0 | 0.00080 | 0.0005 | 0.0012 | 5067 |
| LH Superior Parietal | 3.5002 | 1647.10 | -20.8 | -52.8 | 60.3 | 0.04150 | 0.0390 | 0.0441 | 3811 |
| RH Rostral Middle Frontal | 3.4403 | 6725.81 | 22.9 | 53.1 | 2.5 | < 0.001 | 0.0000 | 0.0002 | 10863 |
| RH Middle Temporal | 2.9419 | 3472.81 | 60.9 | -26.6 | -8.8 | < 0.001 | 0.0000 | 0.0002 | 6476 |
| RH Paracentral | 3.4293 | 1741.04 | 6.1 | -14.5 | 53.0 | 0.02880 | 0.0267 | 0.0310 | 4110 |

*Abbreviations.* LH, left hemisphere; RH, right hemisphere.

**Figure 2.**
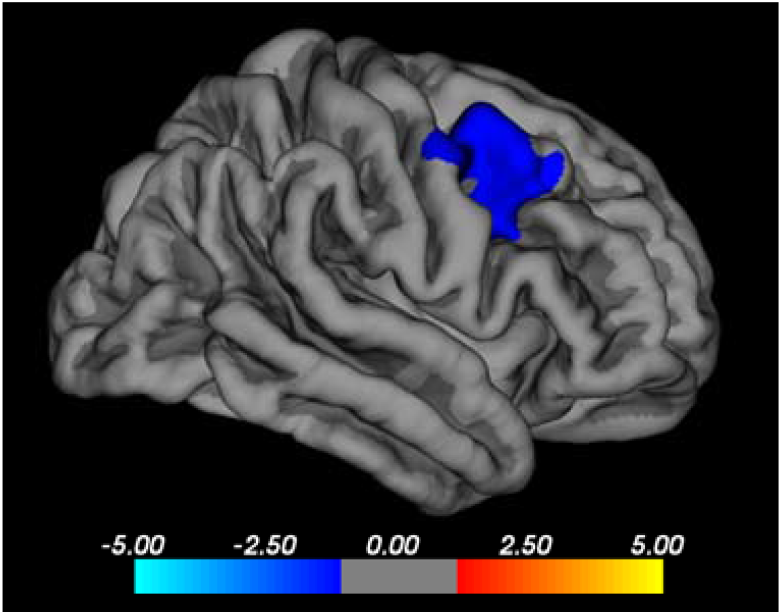
Right hemispheric regional cortical surface area in relation to hyperactivity/ inattention, accounting for gestational age group. Negative correlation between right hemispheric cortical surface area in caudal middle frontal region in relation to hyperactivity/inattention (FDR-corrected *q* < .05).B

Likewise, cortical surface area of the left lingual and inferior parietal regions (Figure 3A), as well as the right superior parietal cluster (Figure 3B) was positively correlated with working memory, indicating that increased surface area was associated with better performance on the WISC-IV Spatial Span test, accounting for gestational age group (LP versus FT) (FDR-corrected *q* < 0.05; Table 3).

**Figure 3.**
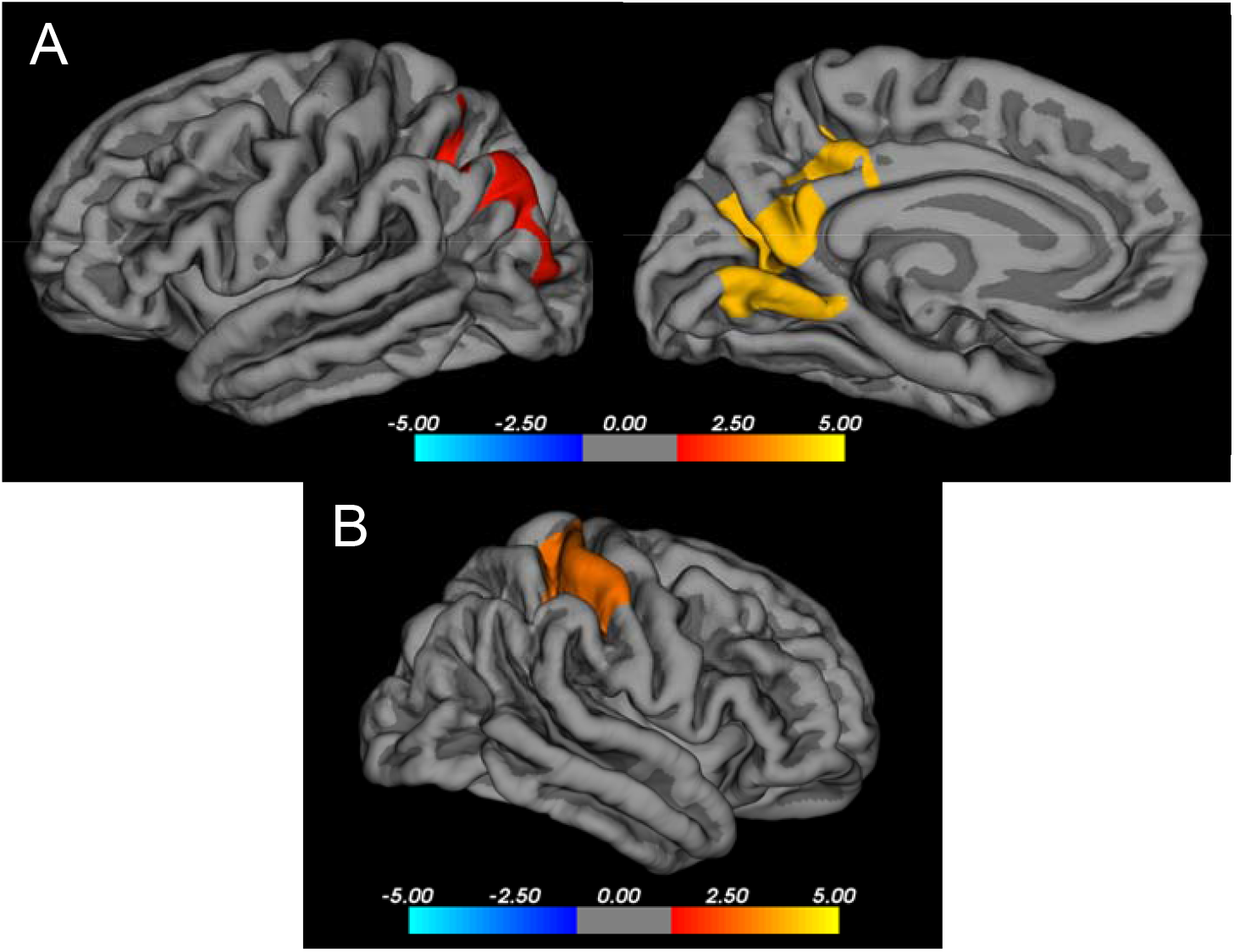
Left (A) and right (B) hemispheric regional cortical surface area in relation to working memory, accounting for gestational age group. Positive correlation between cortical surface area in the left lingual and inferior parietal regions (A) and the right superior parietal cluster (B) in relation to working memory (FDR-corrected *q* < .05).

Lastly, cortical surface area of the left superior frontal, fusiform, and superior parietal regions (Figure 4A) as well as the right rostral middle frontal, middle temporal, and paracentral clusters (Figure 4B) were significantly associated with visual-spatial perception, where increased surface area in both hemispheres was linked to better Benton JLO scores, accounting for gestational age group (LP versus FT) (FDR-corrected q < 0.05; Table 3). No other regional cortical surface area measures were significantly associated with neurocognitive performance after accounting for gestational age group (LP versus FT).

**Figure 4.**
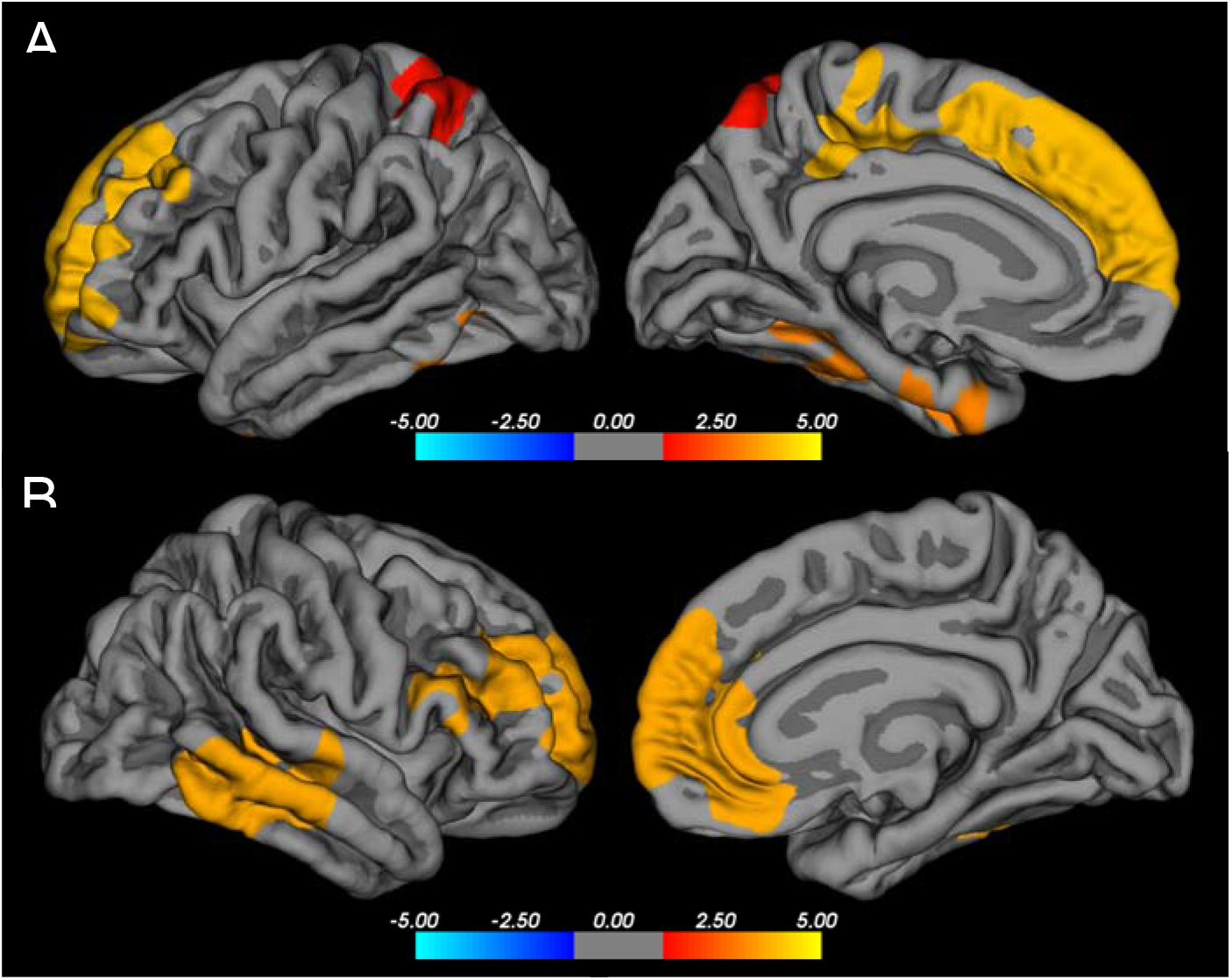
Left (A) and right (B) hemispheric regional cortical surface area in relation to visual-spatial perception, accounting for gestational age group. Positive correlations between cortical surface area in the left superior frontal, fusiform, and superior parietal regions (A) and the right rostral middle frontal, middle temporal, and paracentral clusters (B) in relation to visual-spatial perception (FDR-corrected *q* < .05).

### ROI Analysis of Thalamic Nuclei Across Gestational Age and Neurocognitive Functioning

No comparisons between LP and FT children in thalamic nuclei volumes were statistically significant after correction for multiple comparisons (all FDR-adjusted *q*-values > 0.05). Notably, thalamic nuclei volumes showed bidirectional uncorrected group differences with certain nuclei larger and others smaller in LP children, indicating additional investigations may be warranted in the future (Supplemental Table 1). Likewise, thalamic nuclei volumes were not associated with any of the neurocognitive measures, after accounting for gestational age group (LP versus FT) (all FDR-adjusted *q*-values > 0.05). A small number of main effects reached significance at the uncorrected level; however, none survived correction for multiple comparisons.

## Discussion

The current study utilized complementary MRI approaches – a vertex-wise analysis of cortical surface area and an ROI volumetric analysis of thalamic nuclei – to examine structural morphology between LP and FT children aged 6 to 13 years, and the extent to which structural variations in surface area and thalamic nuclei, more broadly, explain heterogeneity in neurocognitive functioning (hyperactivity/inattention, processing speed, working memory, and visual-spatial perception). Vertex-wise analyses revealed regional differences in cortical surface area between LP and FT children in both hemispheres, along with associations between surface area in several cortical regions and hyperactivity/inattention, working memory, and visual-spatial perception. In contrast, using an ROI approach, thalamic nuclei volumes did not differ between gestational age groups and were not associated with any neurocognitive domains.

These findings extend the work of Brumbaugh et al. 2016 by evaluating regional surface area and thalamic nuclei differences between FT and LP children using FreeSurfer v7.4.1. Notably, vertex-wise analyses revealed differences in cortical surface area of the left lateral orbitofrontal region, as well as right hemispheric differences in the lateral occipital, pars opercularis, inferior temporal, and precentral regions, where LP children exhibited significantly larger surface area compared to the FT counterparts. However, LP children also exhibited lower cortical surface area in the right banks of superior temporal sulcus compared to their FT peers, after correction for multiple comparisons. Furthermore, uncorrected differences in mediodorsal thalamic nuclei (right MDm and right MDl), lateral geniculate nucleus (LGN), pulvinar medial (PuM), and whole right thalamus were directionally consistent with the previously reported whole-thalamus finding, though these did not survive correction for multiple comparisons across nuclei, likely reflecting the increased stringency of subdivided analysis relative to the whole-structure approach used previously. A recent study of 249 neonates found, at 38-44 weeks postmenstrual age, moderate and LP neonates born between 32 0/7 and 36 6/7 weeks of gestation (< 5% had any sort of signal intensity abnormalities, as indicated by hyperintensities on T1-weighted and hypointensities on T2-weighted images, and/or cysts) had smaller corpus callosum, basal ganglia, thalami, and cerebellum volumes, as well as increased extracerebral space, compared to FT neonates.^31^ Similarly, another study of LP neonates (N=29) without intracranial diseases (e.g., hydrocephalus, intraventricular hemorrhage, periventricular leukomalacia) found smaller grey matter volumes, compared to FT neonates at a mean postmenstrual age of 37.5 ± 0.8 weeks, but not in total cerebral volumes or white matter volumes.^30^ In contrast to the current study, which applied stringent exclusion criteria beyond neurological factors, including major medical comorbidities, these more pronounced findings may reflect heightened vulnerability during the early postnatal period, when LP neonates may be exposed to additional physiological stressors (e.g., hypoglycemia).^43,44^

A follow-up study by Rogers et al. 2014 of school-aged 6-to 12-year-olds did not find differences in whole brain, total white matter, hippocampal, corpus callosum, or amygdala volumes between LP and FT children, with LP children having less than 1% difference in total grey matter volume. Exploratory analyses identified smaller right temporal lobes in LP children (superior temporal gyrus and fusiform gyrus), although those effects did not survive Bonferroni correction; reduced right parietal lobe volume (supramarginal gyrus) remained significant following multiple-comparison correction,^43^ unlike our findings which demonstrated larger cortical surface area in the right inferior temporal cluster among LP children. The current study’s findings, in conjunction with the aforementioned study, shed light on the possibility that LP birth may not result in single-direction, widespread, or persistent neuroanatomical alterations in middle to late childhood; rather, the pattern points towards subtle, localized variation in cortical surface area. For instance, the temporal lobe has been highlighted as a region that is particularly susceptible to the deleterious effects of prematurity, especially in LP children, given that it is one of the last areas of the brain to mature.^45^

While LP children exhibited higher rates of hyperactivity/inattention, slower processing speed, poorer working memory, and reduced visual-spatial perception, compared to FT children, as indicated in a previous study of this cohort,^35^ these differences were modest (less than half a standard deviation), reflecting subtle shifts rather than pronounced neurocognitive impairments. Regional cortical surface area was meaningfully associated with neurocognitive performance across several domains, independent of gestational age group. Specifically, right hemispheric cortical surface area in the caudal middle frontal region was negatively associated with hyperactivity/inattention, whereas cortical surface area in the left lingual and inferior parietal regions, as well as the right superior parietal cluster, were positively related to working memory. Lastly, the left superior frontal, fusiform, and superior parietal regions, in addition to the right rostral middle frontal, middle temporal, and paracentral clusters were also positively linked to visual-spatial perception, after accounting for gestational age group. Thalamic nuclei volumes, by contrast, showed no robust associations with neurocognitive functioning, and these relations did not vary by gestational age group.

Few studies have examined the association between brain structure, particularly cortical surface and thalamic nuclei, and neurocognitive outcomes in LP children during middle childhood. However, these regional associations are similar to prior work showing that total grey matter and right temporal lobe volumes were significantly related with later anxiety symptoms in school-aged LP children,^43^ and another reporting associations between cortical surface area in the middle frontal gyrus and working memory and fluid intelligence in a non-clinical sample, consistent with a broader parietofrontal network supporting these cognitive functions.^46^ Collectively, these findings suggest that by school age, LP and FT children may exhibit relatively similar cortical and thalamic nuclei macrostructure, with remaining differences characterized by subtle, localized neuroanatomical variation that is not strongly related to gestational age-associated differences in neurocognitive functioning. Compensatory developmental processes or other factors (e.g., access to developmental and educational resources) may mitigate the role of brain structure in developmental sequelae at this stage of development. It is also possible that these differences, while not driven by macrostructural heterogeneity, may be explained by underlying microstructural and/or network-level circuits tied to specific neurocognitive functions. More advanced neuroimaging techniques (e.g., diffusion MRI, resting-state or task-based functional connectivity) may more effectively elucidate the brain-behavior association, directly capturing white matter microstructure and connectivity relevant to neurocognitive functioning.^47–49^

The current study has several strengths, including extending the limited literature examining brain morphology in school-aged LP children. In addition, despite focusing on macroscopic structural data, we conducted advanced segmentation and vertex-wise analyses to address the possibility that null global findings might conceal subtle, region-specific, or individualized differences. However, several limitations warrant consideration and point to opportunities for future directions. First, sample size was moderate, and second, given the small magnitude of observed neurocognitive differences, substantially larger, likely multicenter samples, will be required to reliably detect neurobiological and neurocognitive group differences in this population. In addition, given early childhood is an important and developmentally meaningful window, its cross-sectional nature may encompass actual differences masked by age-related variation. Furthermore, structural measures such as cortical surface area fail to capture measures of white matter microstructure and functional connectivity. This limited our ability to investigate whether the subtle neurocognitive differences observed between LP and FT children may be due to brain maturational changes related to being born LP. It is plausible that while the present study identified subtle, region-specific differences in cortical surface area with no robust group differences in thalamic nuclei volumes, white matter microstructure and the functional neuroarchitecture may differ.

In conclusion, these findings indicate two distinct patterns: LP and FT children exhibited bidirectional left and right hemispheric cortical surface area differences within a limited set of specific regions, while independent of those group-level differences, individual variation in surface area across a broader set of regions was associated with neurocognitive functioning after accounting for gestational age group among 6–13-year-olds. These findings suggest that LP birth may be linked to subtle, region-specific cortical alterations rather than widespread structural differences during middle childhood. Notably, although regional cortical surface area was associated with several neurocognitive domains across the cohort, these associations did not appear to account for the modest neurocognitive differences associated with LP birth. Future longitudinal research incorporating larger sample sizes, capturing medical risk factors, and utilizing advanced neuroimaging techniques may be needed to better delineate how regional brain differences and brain-behavior associations unfold over time.

## Data Availability

All data produced in the present study are available upon reasonable request to the authors.

