## Supplemental Table 1 for "Cortical surface area, thalamic nuclei volume, and associations with neurocognitive functioning in 6–13-year-old children born late preterm and full term"

**Supplemental Materials**

**Table 1.** Comparison of intracranial volume-normalized thalamic nuclei volumes (x10^3^) between full term and late preterm children.

|  | **Full Term (n=72)** | | **Late Preterm (n=48)** | | **Mean Difference 95% CI** | ***p*-value** | **FDR-Corrected**  ***p*-value** |
| --- | --- | --- | --- | --- | --- | --- | --- |
|  | **M (SD)** | **95% CI** | **M (SD)** | **95% CI** |  |  |  |
| LH AV | 0.099 (0.013) | 0.096, 0.102 | 0.101 (0.014) | 0.096, 0.105 | -0.006, 0.004 | 0.656 | 0.971 |
| LH CeM | 0.053 (0.006) | 0.052, 0.055 | 0.053 (0.007) | 0.051, 0.055 | -0.002, 0.002 | 0.955 | 0.997 |
| LH CL | 0.025 (0.004) | 0.024, 0.025 | 0.025 (0.006) | 0.024, 0.027 | -0.003, 0.001 | 0.358 | 0.821 |
| LH CM | 0.175 (0.015) | 0.172, 0.179 | 0.174 (0.013) | 0.170, 0.178 | -0.004, 0.006 | 0.723 | 0.992 |
| LH LD | 0.026 (0.007) | 0.025, 0.028 | 0.026 (0.009) | 0.024, 0.029 | -0.003, 0.003 | 0.895 | 0.997 |
| LH LGN | 0.204 (0.024) | 0.198, 0.210 | 0.203 (0.022) | 0.197, 0.210 | -0.008, 0.009 | 0.839 | 0.992 |
| LH LP | 0.102 (0.012) | 0.100, 0.105 | 0.101 (0.015) | 0.096, 0.105 | -0.003, 0.007 | 0.484 | 0.879 |
| LH L-Sg | 0.019 (0.005) | 0.018, 0.020 | 0.019 (0.004) | 0.018, 0.020 | -0.002, 0.002 | 0.909 | 0.997 |
| LH MDl | 0.228 (0.025) | 0.222, 0.234 | 0.220 (0.028) | 0.212, 0.228 | -0.002, 0.018 | 0.112 | 0.649 |
| LH MDm | 0.635 (0.057) | 0.622, 0.649 | 0.616 (0.061) | 0.598, 0.633 | -0.002, 0.042 | 0.077 | 0.649 |
| LH MGN | 0.084 (0.013) | 0.081, 0.087 | 0.085 (0.011) | 0.082, 0.088 | -0.005, 0.004 | 0.728 | 0.992 |
| LH Pc | 0.003 (0.000) | 0.003, 0.003 | 0.003 (0.000) | 0.003, 0.003 | 0.000, 0.000 | 0.971 | 0.997 |
| LH Pf | 0.038 (0.003) | 0.037, 0.039 | 0.037 (0.003) | 0.036, 0.038 | -0.001, 0.002 | 0.257 | 0.79 |
| LH Pt | 0.005 (0.001) | 0.005, 0.005 | 0.005 (0.000) | 0.005, 0.005 | 0.000, 0.000 | 0.692 | 0.982 |
| LH PuA | 0.173 (0.017) | 0.169, 0.177 | 0.172 (0.017) | 0.167, 0.177 | -0.006, 0.007 | 0.821 | 0.992 |
| LH PuI | 0.179 (0.024) | 0.173, 0.184 | 0.183 (0.026) | 0.176, 0.191 | -0.014, 0.005 | 0.336 | 0.821 |
| LH PuL | 0.122 (0.017) | 0.119, 0.126 | 0.129 (0.019) | 0.124, 0.135 | -0.013, 0.000 | 0.046 | 0.592 |
| LH PuM | 0.834 (0.086) | 0.814, 0.854 | 0.847 (0.095) | 0.819, 0.874 | -0.046, 0.021 | 0.467 | 0.858 |
| LH VA | 0.318 (0.024) | 0.313, 0.324 | 0.320 (0.028) | 0.312, 0.329 | -0.012, 0.008 | 0.658 | 0.971 |
| LH VAmc | 0.025 (0.002) | 0.024, 0.025 | 0.025 (0.002) | 0.024, 0.025 | -0.001, 0.001 | 0.784 | 0.992 |
| LH VLa | 0.457 (0.038) | 0.448, 0.466 | 0.456 (0.033) | 0.446, 0.465 | -0.011, 0.014 | 0.824 | 0.992 |
| LH VLp | 0.592 (0.049) | 0.580, 0.604 | 0.587 (0.036) | 0.576, 0.597 | -0.010, 0.021 | 0.510 | 0.894 |
| LH VM | 0.015 (0.002) | 0.015, 0.016 | 0.015 (0.001) | 0.014, 0.015 | 0.000, 0.001 | 0.499 | 0.894 |
| LH VPL | 0.637 (0.058) | 0.624, 0.651 | 0.621 (0.052) | 0.606, 0.636 | -0.004, 0.036 | 0.115 | 0.649 |
| LH Whole Thalamus | 5.062 (0.347) | 4.980, 5.143 | 5.034 (0.323) | 4.940, 5.128 | -0.095, 0.151 | 0.654 | 0.971 |
| RH AV | 0.106 (0.012) | 0.103, 0.109 | 0.108 (0.013) | 0.104, 0.112 | -0.007, 0.002 | 0.280 | 0.790 |
| RH CeM | 0.055 (0.006) | 0.054, 0.057 | 0.054 (0.006) | 0.052, 0.055 | -0.001, 0.004 | 0.144 | 0.649 |
| RH CL | 0.025 (0.004) | 0.024, 0.026 | 0.025 (0.005) | 0.023, 0.026 | -0.001, 0.002 | 0.769 | 0.992 |
| RH CM | 0.169 (0.015) | 0.166, 0.173 | 0.167 (0.015) | 0.162, 0.171 | -0.003, 0.008 | 0.315 | 0.804 |
| RH LD | 0.027 (0.007) | 0.025, 0.028 | 0.026 (0.008) | 0.024, 0.029 | -0.002, 0.003 | 0.812 | 0.992 |
| RH LGN | 0.199 (0.022) | 0.194, 0.204 | 0.189 (0.027) | 0.181, 0.197 | 0.001, 0.019 | 0.035* | 0.515 |
| RH LP | 0.101 (0.015) | 0.098, 0.105 | 0.099 (0.014) | 0.095, 0.103 | -0.003, 0.008 | 0.423 | 0.821 |
| RH L-Sg | 0.017 (0.004) | 0.016, 0.018 | 0.017 (0.003) | 0.016, 0.018 | -0.001, 0.002 | 0.412 | 0.821 |
| RH MDl | 0.226 (0.026) | 0.220, 0.232 | 0.214 (0.026) | 0.206, 0.221 | 0.003, 0.022 | 0.013* | 0.356 |
| RH MDm | 0.623 (0.064) | 0.608, 0.638 | 0.590 (0.060) | 0.573, 0.607 | 0.010, 0.055 | 0.005** | 0.356 |
| RH MGN | 0.087 (0.014) | 0.084, 0.091 | 0.088 (0.012) | 0.085, 0.091 | -0.005, 0.004 | 0.819 | 0.992 |
| RH Pc | 0.003 (0.000) | 0.003, 0.003 | 0.003 (0.000) | 0.003, 0.003 | 0.000, 0.000 | 0.087 | 0.649 |
| RH Pf | 0.039 (0.004) | 0.038, 0.040 | 0.039 (0.003) | 0.038, 0.040 | -0.001, 0.002 | 0.365 | 0.821 |
| RH Pt | 0.005 (0.000) | 0.005, 0.005 | 0.005 (0.000) | 0.005, 0.005 | 0.000, 0.000 | 0.244 | 0.79 |
| RH PuA | 0.175 (0.017) | 0.171, 0.179 | 0.170 (0.016) | 0.165, 0.175 | -0.001, 0.011 | 0.097 | 0.649 |
| RH PuI | 0.194 (0.026) | 0.187, 0.200 | 0.182 (0.029) | 0.174, 0.190 | 0.001, 0.022 | 0.027 | 0.442 |
| RH PuL | 0.127 (0.016) | 0.123, 0.131 | 0.129 (0.021) | 0.123, 0.135 | -0.009, 0.005 | 0.549 | 0.934 |
| RH PuM | 0.889 (0.087) | 0.869, 0.910 | 0.853 (0.088) | 0.827, 0.878 | 0.004, 0.069 | 0.027* | 0.442 |
| RH VA | 0.305 (0.024) | 0.300, 0.311 | 0.307 (0.024) | 0.300, 0.314 | -0.010, 0.008 | 0.747 | 0.992 |
| RH VAmc | 0.025 (0.002) | 0.025, 0.026 | 0.024 (0.002) | 0.024, 0.025 | 0.000, 0.001 | 0.082 | 0.649 |
| RH VLa | 0.448 (0.035) | 0.440, 0.457 | 0.445 (0.035) | 0.435, 0.455 | -0.009, 0.016 | 0.600 | 0.956 |
| RH VLp | 0.578 (0.044) | 0.568, 0.589 | 0.570 (0.040) | 0.558, 0.581 | -0.007, 0.024 | 0.286 | 0.79 |
| RH VM | 0.015 (0.002) | 0.014, 0.015 | 0.014 (0.001) | 0.014, 0.015 | 0.000, 0.001 | 0.116 | 0.649 |
| RH VPL | 0.618 (0.054) | 0.605, 0.631 | 0.601 (0.048) | 0.587, 0.615 | -0.002, 0.036 | 0.071 | 0.649 |
| RH Whole Thalamus | 5.072 (0.363) | 4.986, 5.157 | 4.931 (0.312) | 4.840, 5.021 | 0.018, 0.264 | 0.025* | 0.442 |

*Note.* M, SD, and 95% CI values are normalized to ICV and scaled x10^3^. Mean difference values reflect FT relative to LP children. **p*<0.05, ***p*<0.01.

*Abbreviations.* M, mean; SD, standard deviation; CI, confidence interval; LH, left hemisphere; RH, right hemisphere; AV, anteroventral; CeM, central medial; CL, central lateral; CM, centromedian; LD, laterodorsal; LGN, lateral geniculate nucleus; LP, lateral posterior; L-Sg, limitans-suprageniculate; MDl, mediodorsal lateral; MDm, mediodorsal medial; MGN, medial geniculate nucleus; Pc, paracentral; Pf, parafascicular; Pt, paratenial; PuA, pulvinar anterior; PuI, pulvinar inferior; PuL, pulvinar lateral; PuM, pulvinar medial; VA, ventral anterior; VAmc, ventral anterior magnocellular; VLa, ventral lateral anterior; VLp, ventral lateral posterior; VM, ventromedial; VPL, ventral posterolateral.
